# Cerebral hypoperfusion and altered neuro-cardiorespiratory coupling in atrial fibrillation

**DOI:** 10.64898/2026.07.08.26357304

**Authors:** Suk-tak Chan, Ayman Shaqdan, Leon Ptaszek, David Sosnovik, Lok-yiu Do, Bruce R. Rosen, H. Diana Rosas, Jeremy Ruskin, Kenneth K. Kwong

## Abstract

Atrial fibrillation (AF) is associated with an increased risk of neurological morbidity, yet its impact on cerebral perfusion and neuro-cardiorespiratory regulation remains incompletely understood. We used arterial spin labeling, blood oxygenation level-dependent functional MRI (BOLD-fMRI), and a breath-hold challenge to characterize alterations in 14 AF patients compared with 14 age-matched healthy controls. We also examined the changes after catheter ablation with pulmonary vein isolation (PVI) in a subset of patients. Compared with controls, AF patients exhibited widespread reductions in basal cerebral perfusion, including in brainstem regions involved in cardiorespiratory regulation, and a higher prevalence of periodic breathing during wakeful rest. During breath-hold challenge, the coupling between heart rate and BOLD signal changes (ΔBOLD) was smaller in AF, whereas ΔBOLD coupling with breath-by-breath O_2_-CO_2_ exchange ratio was greater at rest within pontine respiratory centers, indicating altered cardiac and respiratory contributions to cerebral hemodynamic regulation. Follow-up MRI scans 1-6 months after PVI demonstrated that restoration of sinus rhythm was associated with stronger heart rate-ΔBOLD coupling during breath-hold challenge, whereas basal cerebral perfusion showed no significant change. This dissociation suggests distinct temporal responses of neuro-cardiorespiratory coupling and cerebral perfusion after sinus rhythm restoration, while the timing of cerebral perfusion recovery remains unresolved.

## 1 Introduction

Atrial fibrillation (AF) is the most common sustained cardiac arrhythmia which is initiated by irregular electrical activity in the atria that disrupts normal pacemaking and produces rapid, erratic atrial contractions.^1, 2^ It is associated with increased risks of stroke, heart failure, and mortality.^3–8^ Beyond its cardiac manifestations, AF frequently co-occurs with a range of neurological morbidities, including sleep apnea, cognitive impairment, and dementia.^9–14^ These associations have largely been attributed to the physiological alterations, such as reduced cerebral perfusion and oxygenation, resulting from irregular heart rates and reduced cardiac output.^15, 16^ However, direct evidence linking AF to changes in cerebral perfusion^17, 18^ and brain physiology (e.g., cerebral oxygenation)^19^ remains limited.

The maintenance of stable cerebral perfusion depends on coordinated interactions among cardiac output, pulmonary gas exchange, and central regulatory networks, particularly within the brainstem.^20^ In healthy individuals, cerebral hemodynamic fluctuations (CHEf) are dynamically coupled to major cardiorespiratory metrics such as heart rate (HR)^21–23^ and the breath-by-breath O_2_-CO_2_ exchange ratio (bER),^24, 25^ a reciprocal measure of pulmonary gas exchange^26–31^ that reflects integrated ventilation-perfusion dynamics. In AF, irregular cardiac rhythm and ventricular responses are associated with increased variability in heart rate^32, 33^ and altered pulmonary gas exchange.^15, 16, 34–36^ These cardiorespiratory disturbances may challenge the central integration of baroreflex and chemoreflex inputs, and cerebrovascular regulation.

In this study, we first sought to characterize alterations in cerebral perfusion and neuro-cardiorespiratory regulation associated with AF. Using arterial spin labeling (ASL) MRI, we quantified basal cerebral perfusion at rest in a cohort of fourteen patients with AF compared to healthy age-matched controls in sinus rhythm. Using blood oxygenation level-dependent functional MRI (BOLD-fMRI), we also assessed CHEf and their coupling to cardiorespiratory physiological signals during both resting conditions and physiological perturbation. A brief breath-hold challenge was used as the perturbation, as it induces transient, endogenous shifts in O_2_ and CO_2_ that remain within the physiological range of ventilation-perfusion regulation in a steady state.

In order to evaluate how sinus rhythm restoration modulated neuro-cardiorespiratory regulation and cerebral perfusion, we also examined a subset of seven AF patients who later underwent catheter ablation with pulmonary vein isolation (PVI), a procedure that restores sinus rhythm. This within-subject pre-post intervention design allowed us to examine the changes in regulatory coupling in relation to cerebrovascular adaptation following sinus rhythm restoration, while minimizing inter-individual variability related to comorbidities.

By integrating cross-sectional neuroimaging with physiological measurements and a within-subject pre-post intervention design, this work aimed to clarify how AF was associated with altered cerebral perfusion and neuro-cardiorespiratory regulation, and how these processes evolved following sinus rhythm restoration. Understanding the relationship between regulatory coupling and cerebral perfusion provided insight into physiological processes underlying neurological vulnerability in AF.

## 2 Materials and Method

### 2.1 Participants

Patients with recurrent atrial fibrillation (AF) were recruited from the Cardiac Arrhythmia Service at Massachusetts General Hospital (MGH). Eligible participants had no unstable medical conditions, including congestive heart failure (Class III and IV of New York Heart Association functional classification), significant valvular heart disease, significant left ventricular dysfunction, ventricular arrhythmias, pulmonary hypertension, uncontrolled hypertension or hypotension, renal insufficiency (creatinine>1.5), or significant pulmonary disease. Patients with an implanted pacemaker or internal defibrillator, prior stroke, or dementia were excluded. Fifteen patients with AF were enrolled and underwent MRI scanning; data from one patient were excluded due to poor fMRI quality.

Among the remaining 14 patients, 10 underwent catheter ablation with pulmonary vein isolation (PVI) and were scanned again 1-6 months post-procedure, to determine whether the presence of AF was directly associated with changes in cerebral perfusion and neuro-cardiorespiratory regulation.

Three patients were excluded from post-procedure analyses due to AF recurrence post PVI before the second MRI scan, lack of follow-up imaging, or poor fMRI quality, resulting in 7 complete pre-/post-PVI datasets. On the day of MRI scanning, an electrocardiogram was used to verify the AF status for each patient before they were positioned in the MRI scanner.

Fourteen age-matched healthy control (HC) participants without a history of AF were recruited from the community, and by e-mail or poster placement within the hospital network of MGH. They were screened to exclude a history of head injury, migraine, sleep apnea, neurologic disorders, psychiatric conditions, or substance abuse. Experimental procedures were explained to the subjects, and signed informed consent was obtained prior to their participation in the study. All components of this study were approved by the Human Research Committee at MGH and conducted in accordance with the Declaration of Helsinki.

### 2.2 Clinical assessments

A comprehensive medication and medical history were obtained from all participants, including a history of diabetes, hyperlipidemia, hypertension, obstructive sleep apnea, valvular heart disease, asthma or smoking history. Cognitive status was assessed during the first visit using Montreal Cognitive Assessment (MoCA) to screen for mild cognitive impairment.

### 2.3 MRI data acquisition

MRI scanning was performed in the Athinoula A. Martinos Center for Biomedical Imaging at MGH on a Siemens Prisma 3-Tesla scanner (Siemens Medical, Erlangen, Germany). Participants’ heads were stabilized using foam padding within a standard 64-channel head coil. The following whole brain MRI datasets were acquired on each participant: 1) structural images using standard high-resolution sagittal images acquired with volumetric T1-weighted 3D-MEMPRAGE sequence (TR=2530ms, TE=1.74ms/3.6ms/5.46ms/7.32ms, flip angle=7°, FOV=256×256mm, matrix=256×256, slice thickness=1mm); 2) perfusion images acquired with pulsed ASL (pASL) sequence (TR=3800ms, TE=13ms, flip angle=90°, bolus duration=600ms, inversion time=1800ms, FOV=220mm×220mm, matrix=64×64, slice thickness=4mm, measurements=121) while the participant was at rest; 3) BOLD-fMRI images acquired with echo planar imaging (EPI) sequence (TR=1250ms, TE=30ms, flip angle=90°, FOV=220×220mm, matrix=108×108, slice thickness=2.4mm, slice gap=1mm, measurements=500) while the participant breathed spontaneously at rest and during a breath-hold challenge. Participants were asked to keep their eyes open during the scans at rest. Visual cues displayed on a projection screen were used to pace the breath-hold challenge, with breath-holding epochs of 30 seconds or less, depending on the participant’s ability to hold their breath, followed by a 90-second epoch of spontaneous breathing; breath-holding and spontaneous breathing could readily be differentiated by their respiratory O_2_ and CO_2_ levels.

Physiological changes, including partial pressures of CO_2_ (PCO_2_) and O_2_ (PO_2_), respiration, and heart rate, were measured simultaneously during the MRI acquisition. A small nasal tubing was placed at the participant’s nostril to sample PCO_2_ and PO_2_ via gas analyzers (Capstar-100, Oxystar-100, CWE, Inc., PA, USA) after calibrating to the barometric pressure on the day of MRI scan, correcting for vapor pressure. Respirations were measured using a respiratory bellow connected to a spirometer (FE141, AD Instruments, Inc., CO, USA). Heart rate (HR) was measured using the electrocardiogram (ECG) module of the Siemens physiological monitoring unit. Plethysmography of a pulse oximeter and a piezoelectric finger sensor was also used to support the identification of R peaks on the ECG tracing for HR measurement. All the physiological recordings were synchronized using the timestamps in the physiological recordings and image headers. BOLD-fMRI images and physiological recordings were stored for offline data analysis.

### 2.4 Processing of physiological data

Physiological data from MRI sessions were analyzed using Matlab R2020b (Mathworks, Inc., Natick, MA, USA) as described previously.^25^ HR was calculated as the inverse of the time interval between the peaks of two consecutive R waves in the ECG time series. Technical delays of PCO_2_ and PO_2_ were corrected via cross-correlation with the respiratory bellow signal. End-inspiration (I) and end-expiration (E) were identified on the PO_2_ and PCO_2_ time series, and were verified by the inspiratory and expiratory phases on the respiratory bellow signal. The duration of the breathing cycle, which is known as the time of breath (ToB), was computed as the time interval between consecutive end-expiration markers. The breath-by-breath end-tidal partial pressures of CO_2_ (P_ET_CO_2_) and O_2_ (P_ET_O_2_) were extracted at each end-expiration. Breath-by-breath O_2_-CO_2_ exchange ratio (bER) was defined as the ratio of the change in PO_2_ to the change in PCO_2_ measured between end-inspiration and end-expiration for each respiratory cycle, the reciprocal of the respiratory gas exchange ratio in the alveolar gas equation.^30^

### 2.5 Preprocessing of pASL data

pASL data were imported into the software Analysis of Functional NeuroImage (AFNI)^37^ (National Institute of Mental Health; http://afni.nimh.nih.gov). After motion correction, voxels located within the ventricles and outside the brain and brainstem identified using FreeSurfer version 7.4^38, 39^ (MGH/MIT/HMS Athinoula A. Martinos Center for Biomedical Imaging, Boston; http://surfer.nmr.mgh.harvard.edu) were excluded from the pASL image analysis. The differences between tag and control images were calculated in pairs using surround subtraction^40^ to minimize contamination from BOLD effects. Longitudinal (T1) relaxation due to the slice-dependent transit delay was compensated based on the acquisition time per slice. pASL volumes were then averaged across all frames and datasets to maximize signal-to-noise. Quantitative maps of cerebral blood flow (CBF) were obtained according to the single-compartment standard kinetic model.^41, 42^ The equilibrium arterial blood magnetization was computed as the intensity in the first scan adjusted for longitudinal (T1) and transverse relaxation (T2*) differences as well as blood-tissue water partition coefficient (λ). Typical values for proton density, λ, T1 and T2* were assumed for all gray matter based on prior literature.^43^ The labeling efficiency was assumed to be 98%;^44^ the mean CBF estimation reliability was greater than 93% with no significant variation across cortical and subcortical structures. Individual participant brain volume with CBF values was registered to the anatomical scan of each participant.

### 2.6 Preprocessing of fMRI data

fMRI data were imported into AFNI version 22.1.14 (Analysis of Functional NeuroImage, NIH; http://afni.nimh.nih.gov)^37^ for preprocessing. The first 12 volumes of each functional dataset before reaching magnetization equilibrium were discarded. Each functional dataset was corrected for artifactual spikes and slice timing. The corrected functional dataset was co-registered to the first image volume using three-dimensional volume registration. The time series for each voxel was normalized to its mean intensity and detrended with fifth-order polynomials to remove low-frequency drift. The components of motion in the time series were removed using linear regression. Motion outliers were defined as any timepoint with 5% or more of brain voxels having an averaged derivative change in translational and rotational motion parameters greater than 0.4.

### 2.7 Correlation analysis for neuro-cardiorespiratory coupling

Individual participant image volumes with the time series of percent BOLD signal changes (ΔBOLD) in the brain were extracted. A Hilbert Transform analysis^45^ was used to measure the cross-correlation between the ΔBOLD and the reference time series of HR and bER for each voxel. Fisher Z-transformation was used to convert the cross-correlation coefficients to Fisher’s z scores.

Individual participant image volumes with Fisher’s z scores were then registered onto the participants’ anatomical scans and transformed to the standardized space of Talairach and Tournoux^46^ for group analysis.

### 2.8 Statistical analysis

Continuous variables (e.g., age) were summarized as median, interquartile range, and coefficient of variation (CoV). The group differences between AF patients and HC were assessed using the Kruskal-Wallis tests. Categorical variables (e.g., sex) were summarized as frequencies; group differences were assessed using the Chi-square tests.

Unpaired t-tests (3ttest++) in AFNI were used to compare the basal cerebral perfusion between AF patients and HC. Unpaired t-tests (3dttest++) in AFNI were also used to compare the HR-ΔBOLD and bER-ΔBOLD correlation coefficients (i.e., Fisher transformed z scores from Hilbert Transform analysis) at rest or under breath-hold challenge between AF patients and HC. The same statistical analysis approach was also applied to compare the basal cerebral perfusion, HR-ΔBOLD and bER-ΔBOLD correlation coefficients measured before and after PVI in AF patients who underwent the procedure.

Monte Carlo simulation was used to correct for multiple comparisons.^47^ Noise smoothness values for the whole brain volume were estimated using the spatial autocorrelation function, and these values were incorporated into a cluster-size threshold simulation program (3dClustSim)^48^ to estimate the probability of false positive clusters via Monte Carlo simulation with 2000 iterations and output a list of corrected thresholds with the corresponding cluster-sizes for the given voxel-wise p-value thresholds. To protect against type I error, a combination of an individual voxel probability threshold of p<0.05, and an estimated contiguous volume was held to correct the overall significance level to α<0.05.

As the cardiorespiratory control networks are mainly located in the brainstem, a small volume correction for multiple comparisons was also performed on the brainstem volume including midbrain, pons, and medulla, and the corrected threshold values with the corresponding cluster-sizes for the given voxel-wise p-value thresholds were estimated. To protect against type I error, a combination of an individual voxel probability threshold of p<0.05, and an estimated contiguous volume was held to correct the overall significance level to α<0.05. To further examine communication between the brainstem and cerebral regions after small volume correction, seed-based connectivity analysis was performed using the brainstem regions identified with significant differences in HR-ΔBOLD and bER-ΔBOLD correlation. Detailed preprocessing and statistical procedures for the seed-based connectivity analysis are provided in the Supplemental Information.

## 3 Results

Fourteen AF patients (median age, 64.5 years; interquartile range IQR, 56.8-69.5 years; 12M) and fourteen HC participants (median [IQR], 62.5 [57.0-67.0] years; 8M) were included for comparison. Several AF patients had known cardiovascular risk factors: five patients had a history of hyperlipidemia, well controlled on statins; nine had a history of hypertension, adequately controlled on anti-hypertensive medications; one had diabetes. Two were prior smokers; four were active smokers. None met the criteria for cognitive impairment based on the performance in the Montreal Cognitive Assessment. No significant differences in age (p=0.71) or gender (p=0.21) were observed between groups (i.e., AF patients and HC). Post-PVI data from 7 AF patients who underwent PVI were included to compare with their pre-PVI data.

### 3.1 Reduced basal cerebral perfusion in AF

Baseline pASL data from 13 AF patients and 12 HC were included in this analysis; pASL data from one patient and two HC were excluded because of motion artifacts. Compared with age-matched HC, AF patients exhibited significantly lower basal cerebral perfusion across widespread cortical (including insula, hippocampus, medial frontal, posteromedial parietal, and occipital cortices), subcortical (including striatum and thalamus), and brainstem structures (including midbrain, dorsal pons, ventrolateral/dorsal medulla) implicated in cardiorespiratory regulation (cluster-corrected p<0.05; Figures 1A and 2).

**Figure 1.**
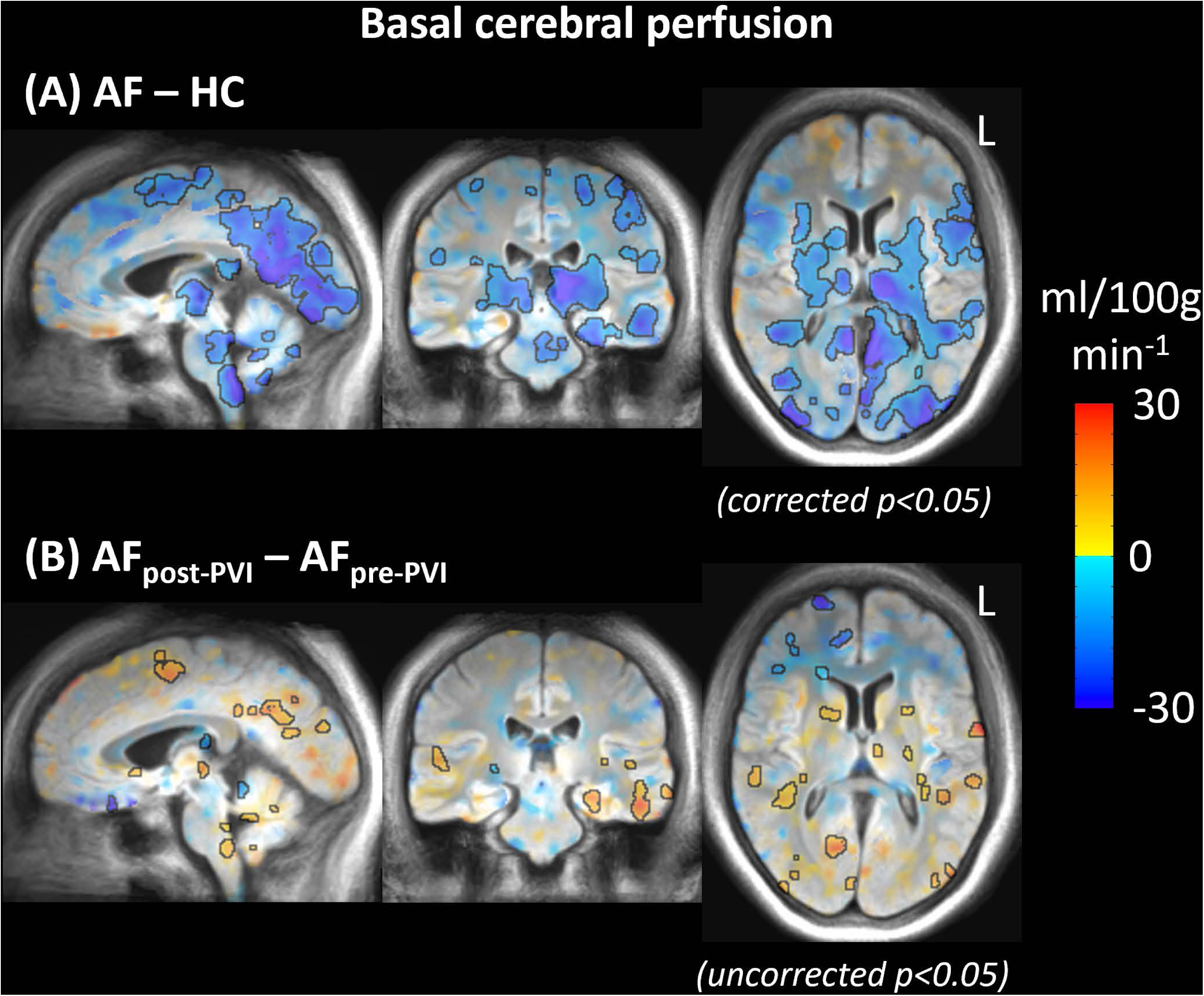
(**A**) Comparison of basal cerebral perfusion between AF patients (n=13) and HC (n=12). Basal cerebral perfusion was significantly lower in AF patients than in HC in the insula, hippocampus, medial frontal cortex, posteromedial parietal cortex (including precuneus and posterior cingulate), occipital cortex, striatum (including caudate nucleus, putamen, and pallidum), thalamus, midbrain (including red nucleus and substantia nigra), dorsal pons, and ventrolateral/dorsal medulla. Cooler colors (cyan to dark blue) on the color bar represent lower basal cerebral perfusion in AF patients relative to HC. Regions outlined in black achieved clustered-corrected p<0.05. (**B**) Comparison of basal cerebral perfusion between pre-PVI scans and post-PVI scans in AF patients (n=7). Warmer colors (red to yellow) indicate greater basal cerebral perfusion following PVI. Regions outlined in black achieved p<0.05 without correction for multiple comparisons.

**Figure 2.**
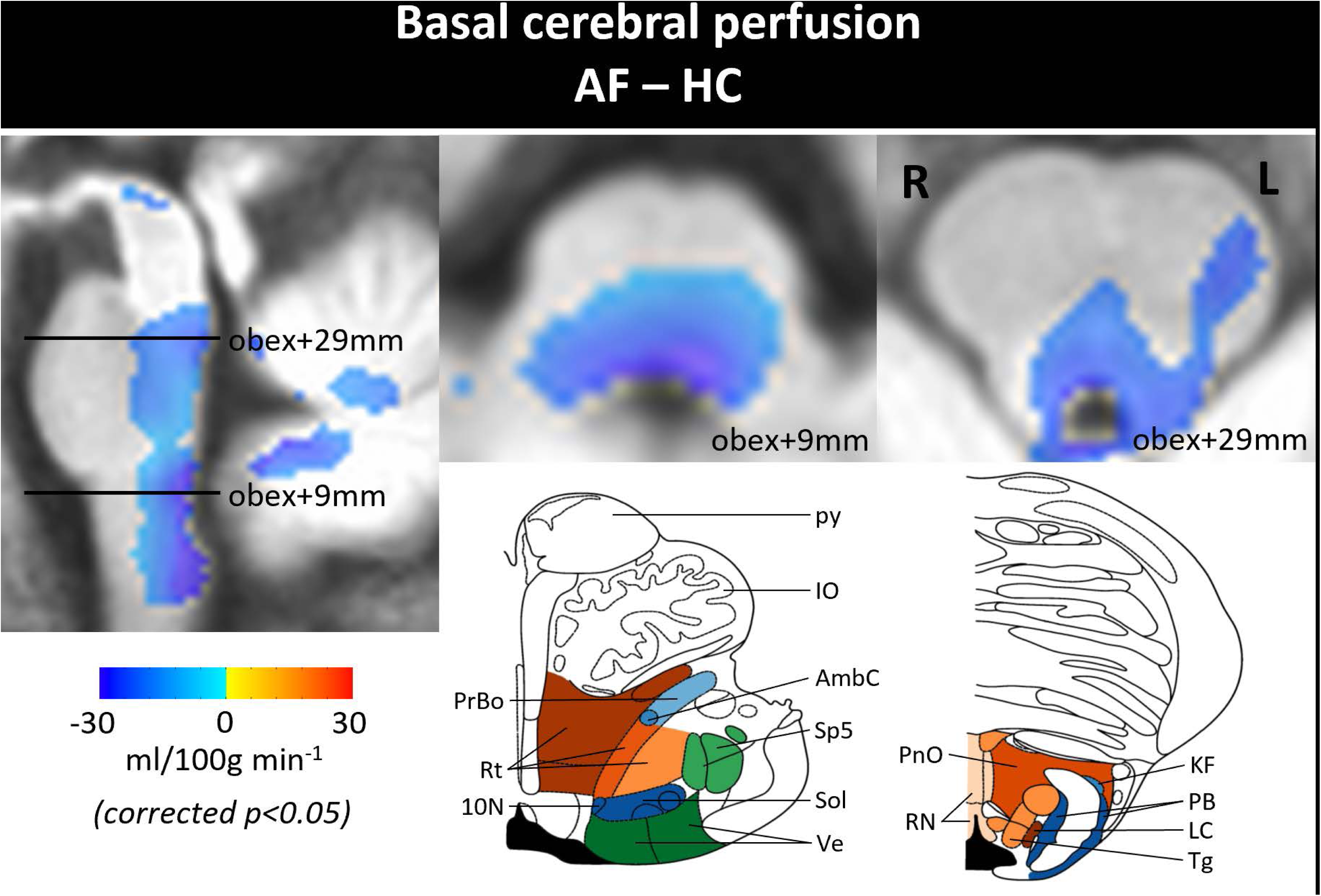
Comparison of basal perfusion in the brainstem between AF patients (n=13) and HC (n=12). Brainstem regions shown in cooler colors (cyan to dark blue) exhibited significantly lower basal perfusion in AF patients compared with HC. Regions shown in colors achieved clustered-corrected p<0.05. Line drawings of the corresponding axial slices at 9mm and 29mm above the obex adapted from Paxinos et al.^85^ are shown in the bottom row. The fourth ventricle is shaded in black. 10N, dorsal nucleus of vagus; AmbC, ambiguous nucleus; IO, inferior olive; KF, Kolliker-Fuse nucleus; LC, locus coeruleus; PB, parabrachial nucleus; PnO, pontine reticular nucleus; PrBo, pre-Bötzinger complex; RN, raphae nuclei; py, pyramidal tract; Rt, reticular nucleus; Sol, nucleus of solitary tract; Sp5, spinal trigeminal tract; Tg, Tegmental nuclei; Ve, vestibular nucleus.

In the subset of AF patients who underwent follow-up MRI scans after PVI, a trend toward increased basal cerebral perfusion was observed in selected brainstem (dorsal pons and medulla) and cortical regions (medial frontal and posteromedial parietal cortex) relative to pre-PVI measurements.

However, these changes did not reach statistical significance after correction for multiple comparisons (Figure 1B).

### 3.2 Increased heart rate variability and altered pulmonary gas exchange at rest in AF

Physiological metrics measured at rest in AF patients and HC are summarized in Table S1. At rest, AF patients exhibited a higher mean HR compared with HC (AF: 77.53 [67.21-88.24] beats/min; HC: 67.28 [64.23-73.41] beats/min; p=0.027), along with significantly greater HR variability, as reflected by an increased coefficient of variation (AF: 0.18 [0.16-0.20]; HC: 0.05 [0.03-0.06]; p<0.001; Figure S1-A). In contrast, while the mean bER did not differ significantly between groups (AF: 1.17 [1.09-1.26]; HC: 1.12 [1.06-1.25]; p=0.491), variability in bER was significantly greater in AF patients than in HC (AF: 0.10 [0.09-0.12]; HC: 0.07 [0.05-0.08]; p=0.004). These differences are demonstrated in the resting HR and bER time series from a representative HC (Figure 3A, upper panel) and a representative AF patient (Figures 3B, upper panel).

**Figure 3.**
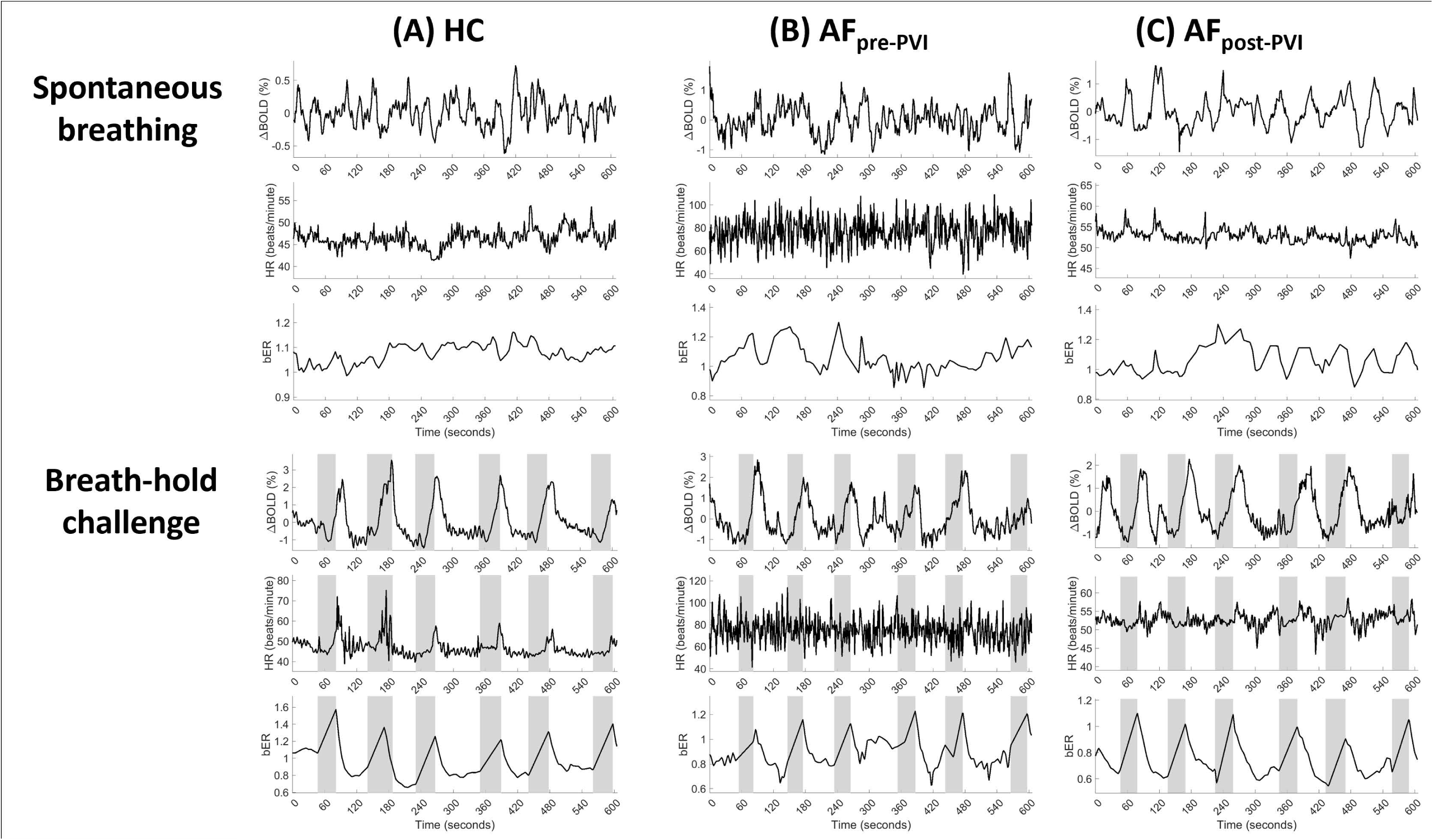
Time series of ΔBOLD extracted from whole-brain gray matter, HR and bER during spontaneous breathing at rest and breath-hold challenge in a representative (**A**) HC, and a representative AF patient (**B**) before PVI, and (**C**) after PVI.

Irregular breathing patterns, including irregular pauses or periodic breathing, were observed more frequently in AF patients than in HC. Periodic breathing was noted in 8 out of 14 AF patients (57%) and in 3 of 14 HC participants (21%). Periodic breathing during awake resting conditions was defined as the presence of at least 3 consecutive cycles of ventilatory variation exceeding 25% of the peak-to-trough amplitude of a typical sinusoidal breathing pattern.^49, 50^ The time series of bER and respiration measured before and after PVI in 2 representative patients are shown in Figure S2. Despite this difference in prevalence, no significant group differences were observed in the mean or variability of time-of-breath between pre- and post-PVI.

In the subset of AF patients who underwent follow-up imaging after PVI, resting mean HR (Pre-PVI: 82.60 [80.50-93.34] beats/min; Post-PVI: 61.69 [53.50-63.53] beats/min; p=0.031) and HR variability were significantly reduced relative to pre-PVI measurements (Pre-PVI: 0.19 [0.17-0.20]; Post-PVI: 0.03 [0.02-0.04]; p=0.016; Figure S1-B). Mean bER was also significantly reduced following PVI (Pre-PVI: 1.21 [1.12-1.28]; Post-PVI: 1.08 [1.05-1.16]; p=0.016), but there was no significant difference in bER variability (Pre-PVI: 0.11 [0.09-0.12]; Post-PVI: 0.09 [0.06-0.10]; p=0.219; Table S2). Resting HR and bER time series from a representative AF before and after PVI are shown in Figures 3B and 3C (upper panels).

### 3.3 Reduced heart-brain coupling during physiological challenge in AF

Time series of ΔBOLD in whole-brain gray matter and the corresponding HR and bER from a representative HC and a representative AF patient during breath-hold challenge are shown in Figures 3A and 3B (lower panels), respectively. In both the AF patient and HC, bER showed temporal correspondence with ΔBOLD during resting conditions and during the breath-hold challenge. In contrast, the AF patient exhibited rapid and irregular HR fluctuations at rest and during breath-holding, whereas HR in HC demonstrated temporal correspondence with ΔBOLD during portions of the time series.

At the group level, cross-correlation analyses revealed no significant difference in HR-ΔBOLD coupling at rest between AF patients and HC. During the breath-hold challenge, however, HR-ΔBOLD coupling was significantly reduced in AF patients across multiple cortical (including medial and ventromedial prefrontal, hippocampal, lateral temporal, posteromedial parietal, and occipital cortices), subcortical (including striatum, basal forebrain, hypothalamus, and thalamus), and brainstem regions (including the midbrain, dorsal and rostral pons, and dorsal medulla) involved in cardiorespiratory regulation (cluster-corrected p<0.05; Figures 4A and 5). Most of these regions overlapped with areas showing reduced basal cerebral perfusion in AF patients (Figures 1A and 2).

**Figure 4.**
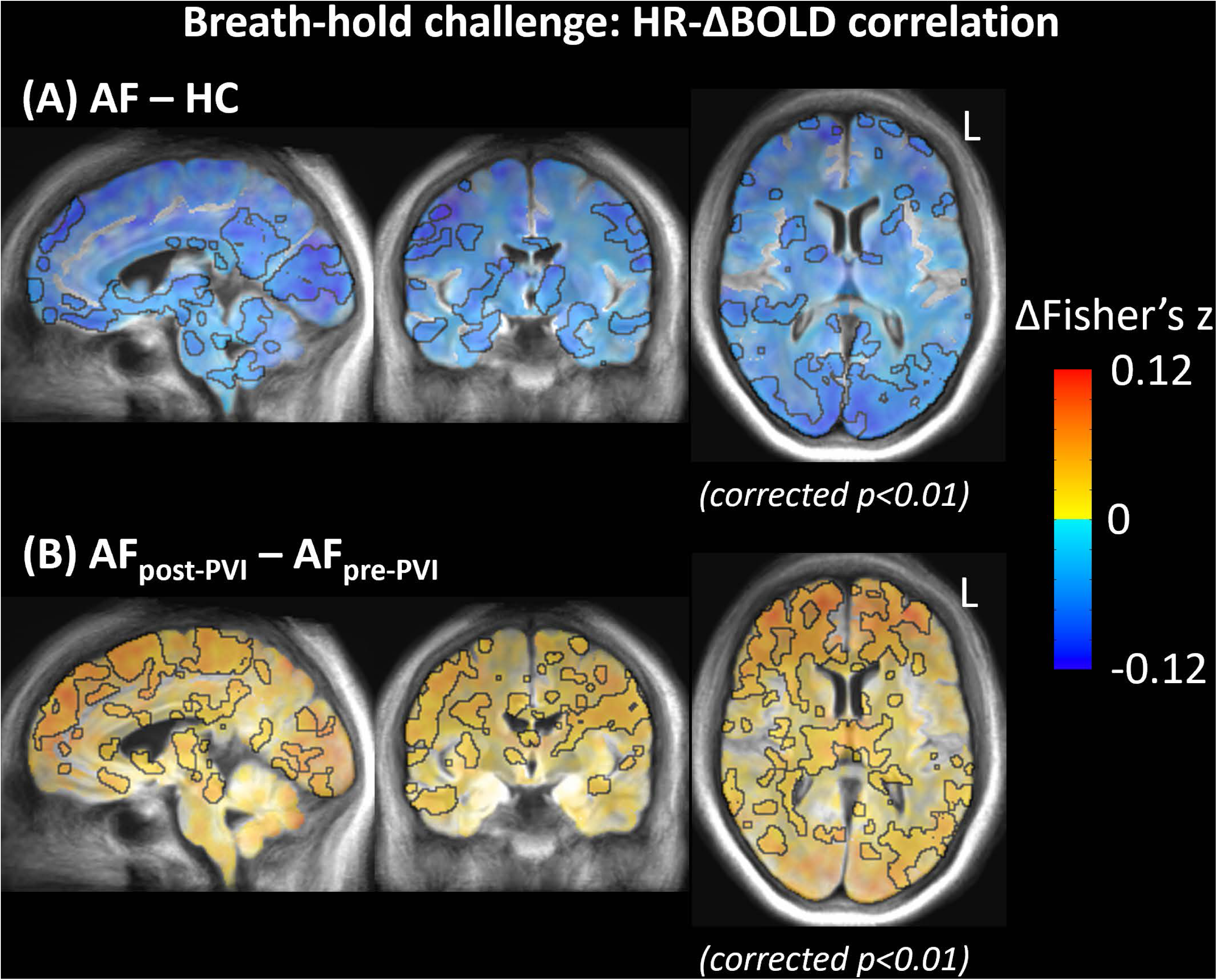
(**A**) Comparison of HR-ΔBOLD correlation between AF patients (n=14) and HC (n=14) during breath-hold challenge. The HR-ΔBOLD correlation was significantly smaller in AF patients than in HC in medial and ventromedial prefrontal cortices, hippocampus, lateral temporal cortex, posteromedial parietal cortex (including precuneus and posterior cingulate), occipital cortex, striatum (including caudate nucleus, putamen, and pallidum), basal forebrain, hypothalamus, thalamus, midbrain (including red nucleus and substantia nigra), rostral and dorsal pons, and ventrolateral/dorsal medulla. Cooler colors (cyan to dark blue) on the color bar represent smaller HR-ΔBOLD correlation in AF patients relative to HC. Regions outlined in black achieved clustered-corrected p<0.05. (**B**) Comparison of HR-ΔBOLD correlation between pre-PVI scans and post-PVI scans in AF patients (n=7) during breath-hold challenge. Warmer colors (red to yellow) indicate greater HR-ΔBOLD correlation following PVI. Regions outlined in black achieved clustered-corrected p<0.05.

**Figure 5.**
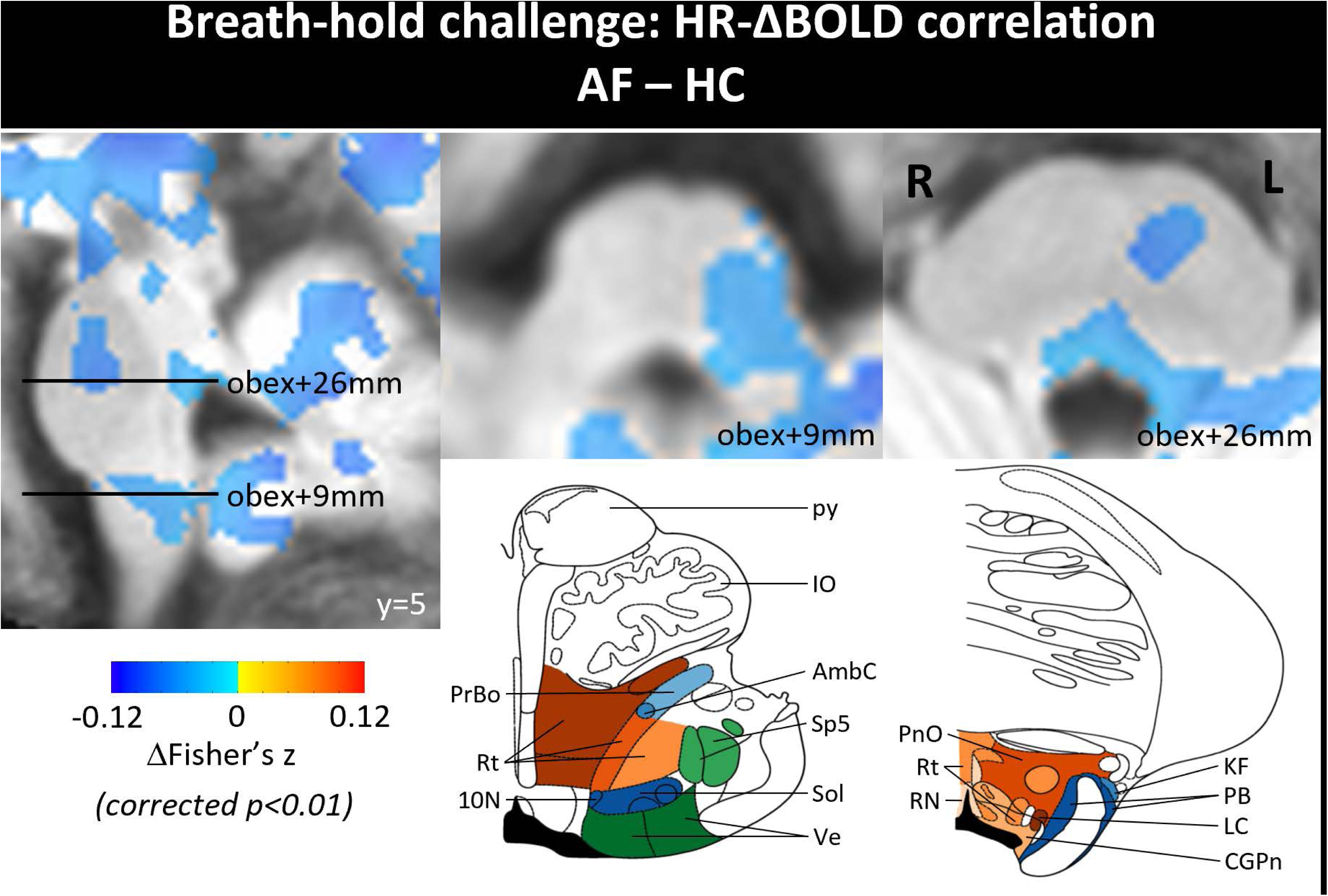
Comparison of HR-ΔBOLD correlation in brainstem between AF patients (n=14) and HC (n=14) during breath-hold challenge. Brainstem regions shown in cooler colors (cyan to dark blue) showed significantly smaller HR-ΔBOLD correlation in AF patients compared with HC. Regions shown in colors achieved clustered-corrected p<0.01. Line drawings of the corresponding axial slices at 9mm and 26mm above the obex adapted from Paxinos et al.^85^ are shown in the bottom row. The fourth ventricle is shaded in black. 10N, dorsal nucleus of vagus; AmbC, ambiguous nucleus; CGPn, central gray pons; IO, inferior olive; KF, Kolliker-Fuse nucleus; LC, locus coeruleus; PB, parabrachial nucleus; PnO, pontine reticular nucleus; PrBo, pre-Bötzinger complex; RN, raphae nuclei; py, pyramidal tract; Rt, reticular nucleus; Sol, nucleus of solitary tract; Sp5, spinal trigeminal tract; Ve, vestibular nucleus.

In the subset of AF patients who underwent follow-up imaging after PVI, HR-ΔBOLD coupling during the breath-hold challenge was significantly greater than in pre-PVI scans (Figure 4B). As expected, HR time series from the same representative AF patient demonstrated that the rapid and irregular HR fluctuations observed at rest or during breath-holding prior to PVI (Figure 3B) were no longer evident following PVI (Figure 3C). HR showed clearer temporal correspondence with ΔBOLD during portions of the post-PVI scan.

### 3.4 Greater lung-brain interactions at rest in AF patients

During spontaneous breathing, bER-ΔBOLD coupling was significantly greater in AF patients than in HC within brainstem regions involved in cardiorespiratory control, including the dorsal and ventral pons (Figure 6) after small volume correction. To further examine the connection between brainstem and cerebral regions, seed-based connectivity analysis was performed using the identified pontine regions showing significant differences in bER-ΔBOLD coupling. Compared with HC, AF patients exhibited significantly greater resting-state functional connectivity between the pons and the left insula (Figure S3).

**Figure 6.**
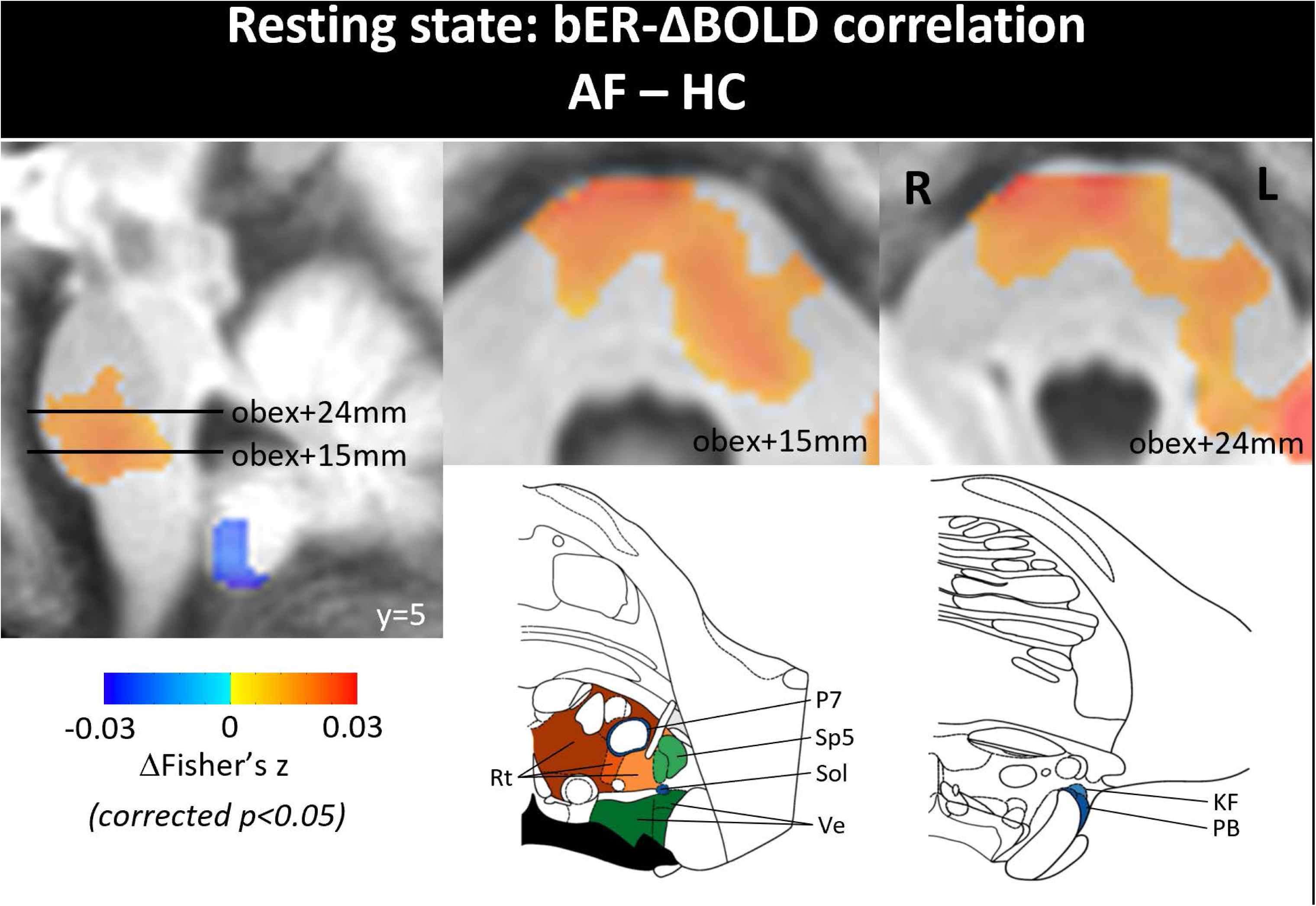
Comparison of bER-ΔBOLD correlation in brainstem between AF patients (n=14) and HC (n=14) at rest. Brainstem regions shown in warmer colors (red to yellow) showed significantly greater bER-ΔBOLD correlation in AF patients compared with HC. Regions shown in colors achieved clustered-corrected p<0.05. Line drawings of the corresponding axial slices at 15mm and 24mm above the obex adapted from Paxinos et al.^85^ are shown in the bottom row. The fourth ventricle is shaded in black. KF, Kolliker-Fuse nucleus; P7, perifacial zone; PB, parabrachial nucleus; Rt, reticular nucleus; Sol, nucleus of solitary tract; Sp5, spinal trigeminal tract; Ve, vestibular nucleus.

During the breath-hold challenge, no significant difference in bER-ΔBOLD coupling was observed between AF patients and HC. In addition, bER-ΔBOLD coupling at rest and during the breath-hold challenge did not show significant change in AF patients before and after PVI.

## 4 Discussion

We found that AF was associated with reduced basal cerebral perfusion and altered neuro-cardiorespiratory coupling. These changes were accompanied by increased HR variability and tachycardia and were observed across cortical, subcortical, and brainstem regions involved in cardiorespiratory regulation. By combining cross-sectional comparisons between AF patients and HC with within-subject pre-post PVI comparisons in AF patients, this study demonstrated that disturbances in cerebral perfusion and regulatory coupling are present in AF, and that these processes exhibited distinct temporal responses following restoration of sinus rhythm. In particular, heart-brain coupling during physiological challenge improved after PVI, while the basal cerebral perfusion showed no significant change over the same time interval, and the timing of cerebral perfusion recovery remains unresolved.

Using an arterial spin labeling technique to quantify regional cerebral perfusion, we observed significantly lower basal perfusion in patients with AF across cortical, subcortical, and brainstem regions. Affected cortical regions included the core brain regions within resting-state networks for different brain functions (e.g., medial frontal and posteromedial parietal cortices in default mode network, hippocampus in limbic network, occipital cortex in visual network, and insula in all these three networks).^51^ Notably, many of these together with the subcortical regions overlap with areas previously associated with gray matter alterations and cognitive decline in AF,^52^ providing a physiological context for neurological vulnerability. In addition, brainstem involvement encompassed medullary and pontine regions overlapping the reticular formation, including the locus coeruleus, as well as the dorsal vagal complex, including dorsal motor nucleus of vagus and nucleus of solitary tract, which play central roles in heart rate modulation and cardiorespiratory integration.^53–55^

Beyond reduced cerebral perfusion, our findings demonstrated that altered heart-brain interactions during physiological challenge (i.e., breath-holding) in patients with AF. Breath-holding has been commonly used as a challenge to induce physiological stress or transient disturbances in ventilation and pulmonary perfusion coordinated by the brainstem cardiorespiratory control centers. In healthy individuals, HR and bER increased during breath-holding, and the corresponding changes in CHEf were observed (Figure 3A), consistent with intact cardiac and respiratory contributions to cerebral hemodynamic regulation. In contrast, in AF patients, HR showed limited modulation during breath-holding despite preserved bER responses, resulting in reduced HR-ΔBOLD coupling across cortical, subcortical, and brainstem regions compared to HC (Figure 3B). These regions include components of the ascending reticular activating system^56, 57^ which are involved in the regulation of arousal and consciousness,^56–58^ and sleep-wake cycle (e.g., reticular formation, thalamus, hypothalamus, basal forebrain),^59^ in addition to the posteromedial parietal, sensorimotor, and visual cortices, highlighting the broad engagement of heart-brain interactions during cardiorespiratory challenge. Restoration of sinus rhythm following PVI was associated with normalization of HR variability and HR-ΔBOLD coupling during breath-holding (Figure 3C). Together, breath-holding revealed exaggerated differences in heart-brain coupling between AF patients and HC, consistent with impaired cardiac modulation of heart rate during cardiorespiratory challenge.

The impact of AF was evident not only during physiological challenge but also during spontaneous breathing at rest. As expected, compared with HC, patients with AF exhibited rapid and irregular HR fluctuations,^1, 2^ reflected by significantly greater HR variability. Greater variability in bER was also observed in AF patients. Such variability may be attributed to alterations in cardiopulmonary dynamics associated with AF, including loss of atrioventricular asynchrony, irregular ventricular responses, and altered pulmonary and ventricular pressure dynamics.^16, 36^ A plausible physiological explanation is that reduced effective cardiac output and associated circulatory delay, together with fluctuations in pulmonary gas exchange, may increase chemoreflex-related ventilatory drive and reduce ventilatory stability, thereby predisposing to periodic breathing even during awake resting conditions.^50, 60^ Consistent with this framework, periodic breathing during awake resting condition was observed more frequently in AF patients than in HC. Prior work has linked daytime periodic breathing to adverse outcomes in patients with heart failure.^49, 61^ Periodic breathing, which persisted in some patients following PVI, suggests that restoring sinus rhythm was not consistently accompanied by normalization of resting breathing patterns.

Additionally, while no significant difference in heart-brain interaction in terms of HR-ΔBOLD coupling was observed at rest between AF patients and HC, altered lung-brain interactions in AF patients during spontaneous breathing were indicated by greater bER-ΔBOLD coupling in AF patients within the ventrolateral and dorsal pons, encompassing structures implicated in respiratory rhythm generation, including post-inspiratory complex and parafacial respiratory group,^62^ as well as components of the pontine noradrenergic system for cardiorespiratory coupling such as the A5 cell group.^62–64^ Area A5 has been associated with modulation of respiratory rhythm and cardiovascular control, particularly under hypoxic or hypercapnic conditions.^63^ Greater bER-ΔBOLD coupling in these regions may therefore reflect increased sensitivity of brainstem cardiorespiratory regulatory networks to breath-by-breath fluctuations in pulmonary gas exchange during spontaneous breathing in AF. Compared with HC, greater bER-ΔBOLD coupling was also observed in the left insula in AF patients at rest. Altered brain connectivity between left insula and pontine regions may reflect shared anatomical and neurochemical features, including the presence of noradrenergic neuron populations,^63–66^ that support coordinated lung-brain interactions during resting condition. Insular involvement has been associated with AF detected after stroke or transient ischemic attack ^67–70^, and experimental models have demonstrated atrial fibrosis, particularly at the left atrium-pulmonary vein border, following insular ischemic stroke.^71^ Although the present cohort consisted of patients with AF of cardiogenic origin, the observed insular involvement highlights a potential brain-heart interaction pathway in cardiogenic AF that warrants further investigation in future mechanistic and longitudinal studies.

Catheter ablation with PVI can restore sinus rhythm and reduce HR variability; however, available evidence suggests that cerebral perfusion does not improve consistently in patients with paroxysmal AF. Takahashi et al.^72^ reported increased blood flow in the internal carotid and basilar arteries six months after PVI in patients with non-paroxysmal AF, but not in those with paroxysmal AF. Similarly, Hashimoto et al.^73^ observed increases in cerebral and hippocampal blood flow three months after PVI in patients with non-paroxysmal AF, whereas no comparable improvement was detected in patients with paroxysmal AF. These prior observations are consistent with our finding that regional cerebral perfusion, measured by ASL, did not change significantly on follow-up MRI scans obtained 1 to 6 months after PVI. While the timing and the extent of cerebral perfusion recovery remains unresolved, the absence of a significant perfusion response during follow-up, together with prior reports of persistent affective, cognitive, and physical symptoms after ablation,^74–78^ underscores the need for longer-term studies linking cerebral perfusion with clinical outcomes.

In contrast, HR-ΔBOLD coupling during breath-hold challenge increased significantly across multiple cerebral regions following PVI, consistent with improved heart-brain regulatory coupling after sinus rhythm restoration (Figure 4B). However, several regions associated with cholinergic and noradrenergic effects which could play a role on cardiac autonomic regulation reflected in HR variability,^79^ including the reticular formation, hippocampus, basal forebrain,^80^ precuneus, lateral occipital cortex, locus coeruleus,^81, 82^ and nucleus of solitary tract, did not show significant post-PVI changes in HR-ΔBOLD coupling (Figure 4B). The lack of significant post-PVI changes in these cerebral and brainstem regions, together with the marked reduction in HR variability to levels lower than those observed in HC, is consistent with incomplete normalization of cardiac autonomic modulation after PVI, potentially reflecting ablation-related parasympathetic denervation.^83, 84^

## 5 Limitations

Several limitations of our study should be considered. The sample size of this AF cohort was relatively small. In addition, only a subset of AF patients underwent PVI based on clinical indications, limiting statistical power for the analyses of pre-post PVI changes.

Because AF predominantly affects older adults, age-related alterations in cerebral perfusion and neuro-cardiorespiratory coupling may partially obscure disease-related effects. To mitigate this, we included age-matched healthy participants for comparison. Some AF patients had hypertension and diabetes that were controlled on medications. To minimize inter-individual variability related to comorbidities, we included a within-subject comparison of measurements before and after PVI. Quantification of heart rate during MRI in patients with AF can also be challenging due to inherent rhythm irregularity. To address this, ECG was complemented with optical and piezoelectric plethysmography to support robust heart rate estimation.

Finally, given the distinct temporal responses of neuro-cardiorespiratory coupling and cerebral perfusion observed after PVI, longer-term longitudinal studies, including follow-up beyond one year, may be necessary to more fully characterize cerebral perfusion recovery and the evolution of brain-heart-lung interactions following sinus rhythm restoration.

## 6 Conclusion

Our findings indicate that AF is associated with reduced basal cerebral perfusion, an increased prevalence of periodic breathing during wakeful rest, decreased heart-brain coupling during physiological challenge, and increased lung-brain coupling at rest compared with healthy controls. Together, these findings support a model in which AF disrupts both cerebral perfusion and the balance of cardiac and respiratory contributions to cerebral hemodynamic regulation. Restoration of sinus rhythm following PVI was associated with stronger heart-brain coupling during the breath-hold challenge, whereas cerebral perfusion showed no significant change over the same follow-up interval. This dissociation suggests distinct temporal responses of neuro-cardiorespiratory coupling and cerebral perfusion following sinus rhythm restoration, while leaving the relative timing and extent of cerebral perfusion recovery unresolved. Future studies with larger cohorts and longitudinal follow-up, including correlations with clinical and cognitive outcomes, may help to determine whether and how cerebral perfusion changes over time, and to further clarify the physiological significance of altered brain-heart-lung interactions in AF and their potential relevance to neurological vulnerability.

## Acknowledgments

The authors gratefully acknowledge all volunteers for their time and participation in this study.

## 7 Author Contribution Statement

STC, DS, JR and KKK idea conception; STC, AS, LP and KKK data collection; STC, AS and KKK data analysis; STC, AS, LYD, KKK figure preparation and draft of manuscript; STC, AS, LP, DS, BRR, HDR, JR and KKK revision, critique and final manuscript approval.

## 8 Declaration of Conflicting Interest

Dr. Ruskin has served as a consultant for Acesion Pharma, Advanced Medical Education, Alnylam, Baim Institute/Lexeo, InCarda, Janssen, Sanofi, Treeline Bio, and Vertex and has equity in Amgen, Celero Systems, Element Science, Infobionic, and Regeneron.

## 9 Ethical Approval

All components of this study were approved by the Human Research Committee at Massachusetts General Hospital (2013P001047).

## 10 Funding

This research was performed in whole at the Athinoula A. Martinos Center for Biomedical Imaging at the Massachusetts General Hospital, using resources provided by the Center for Functional Neuroimaging Technologies, P41EB015896 (BRR), a P41 Biotechnology Resource Grant supported by the National Institute of Biomedical Imaging and Bioengineering (NIBIB), National Institutes of Health, as well as the Shared Instrumentation grants S10RR023043 (BRR) and S10OD034213 (KKK). This study was also supported, in part, by National Center for Complementary and Integrative Health (NCCIH) grants R21AT010955 (KKK) and R21AT012386 (KKK, STC), National Institute for Neurological Disorders and Stroke grants R01NS106384 (HDR) and R01NS114562 (HDR).

## 11 Data Availability

All relevant data, not including subject identifiers, are within the manuscript. Individual imaging data with subject identifiers cannot be shared publicly due to institutional policies regarding the protection of research subject privacy; the consent form signed by subjects also did not include language which would permit the inclusion of their images in public data repositories. However, data generated or analyzed during the study are available for researchers who meet criteria established by Massachusetts General Hospital IRB. Researchers seeking to utilize de-identified imaging data from this manuscript should contact the corresponding author (Dr. Suk-tak Chan).

## 12 Supplemental Material

A Word document (SupplInfo.docx) with supplemental text and figures is attached.

