## Supplementary material for "Cerebral hypoperfusion and altered neuro-cardiorespiratory coupling in atrial fibrillation": SupplInfo

**Supplemental Information**

**Title**: Cerebral hypoperfusion and altered neuro-cardiorespiratory coupling in patients with paroxysmal atrial fibrillation

**Authors**: Suk-tak Chan, PhD, Ayman Shaqdan, MD, Leon Ptaszek, MD, PhD, David Sosnovik, MD, Lok-yiu Do, BA, Bruce R. Rosen, MD, PhD, Herminia D. Rosas, MD, Jeremy Ruskin, MD, Kenneth K. Kwong, PhD

**Seed-based resting-state connectivity analysis between brainstem and cerebral regions**

Resting-state functional connectivity analysis was performed to examine communication between the brainstem and cerebral regions. Following motion correction, the preprocessed functional data were temporally band-pass filtered (0.008–0.125 Hz). Voxels within the ventricles and outside the brain, as defined by the FreeSurfer-parcellated brain volume,[1, 2] were excluded from further analysis.

For each participant, image volumes containing time series of percent BOLD signal change (ΔBOLD) were computed. Brainstem regions showing significant differences between AF patients and HC were identified, and the ΔBOLD time series within these regions were averaged to generate a seed signal for connectivity analysis. Pairwise Pearson correlation coefficients were then calculated between the seed time series and the ΔBOLD time series of each voxel across the brain using '*3dNetCorr*'[3] in AFNI. Correlation coefficients were converted to Fisher's $z$ scores for subsequent group-level analyses.

Group comparisons of Fisher-transformed z scores were conducted using unpaired t-tests (3dttest++) in AFNI, comparing AF patients with HC. Correction for multiple comparisons was performed using Monte Carlo simulation.[4] Spatial noise smoothness for the whole brain volume was estimated using the spatial autocorrelation function, and these estimates were incorporated into *3dClustSim*[5] to generate cluster-size thresholds based on 2000 simulations. To protect against type I error, a combination of an individual voxel probability threshold of $p<0.05$, and an estimated contiguous volume was held to correct the overall significance level to $\alpha<0.05$.

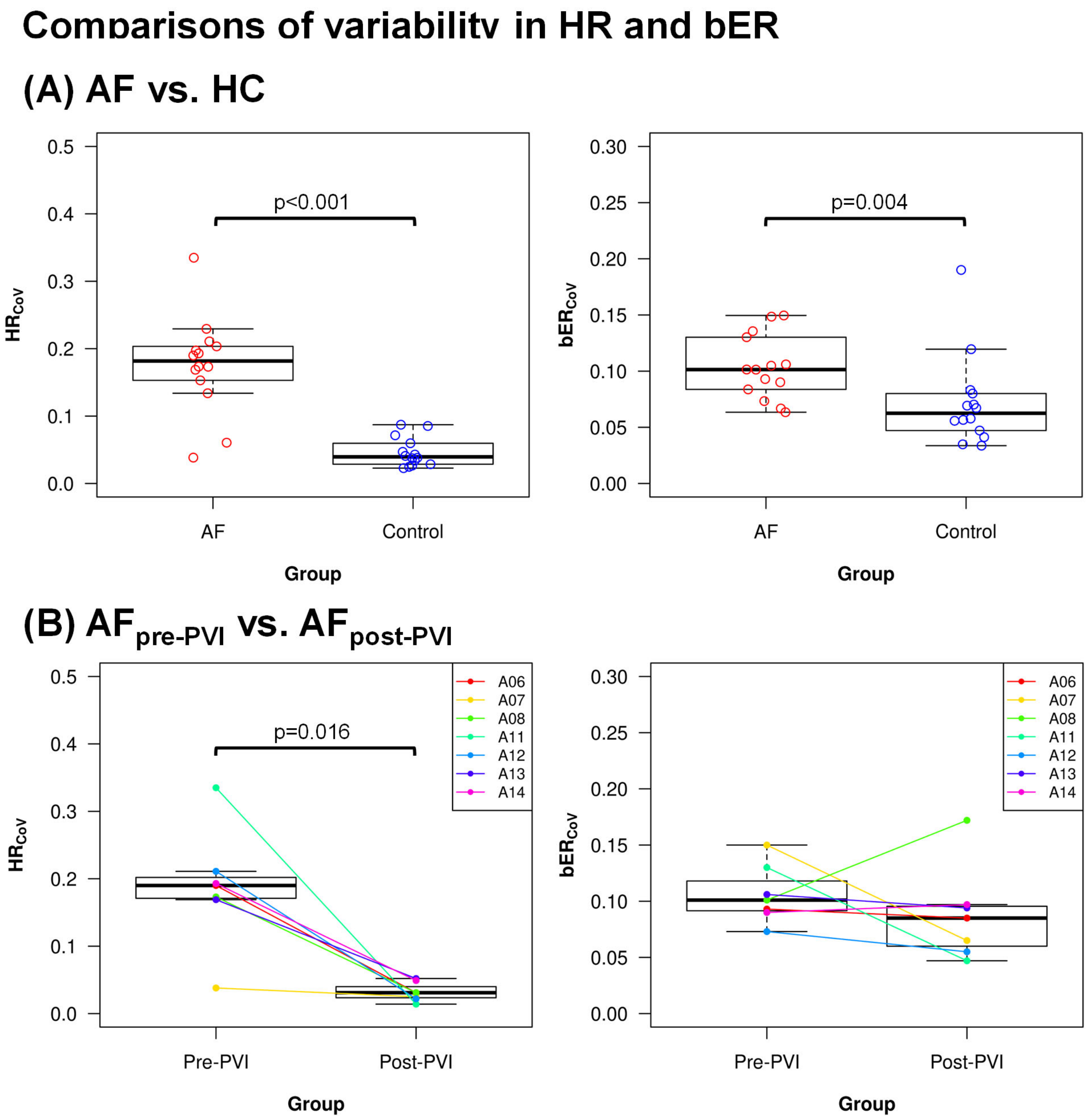


**Figure S1**. Comparison of variability in HR and bER **(A)** between AF patients before PVI (n=14) and HC (n=14), and **(B)** between pre-PVI scans ($AF_{pre\text{-}PVI}$) and post-PVI scans ($AF_{post\text{-}PVI}$) in AF patients (n=7). Variability in both HR and bER were significantly greater in AF patients than in HC. HR variability was significantly reduced after PVI compared with pre-PVI scans, whereas bER variability did not change significantly. Variability is expressed as the coefficient of variation in HR ($HR_{CoV}$) and bER ($bER_{CoV}$).

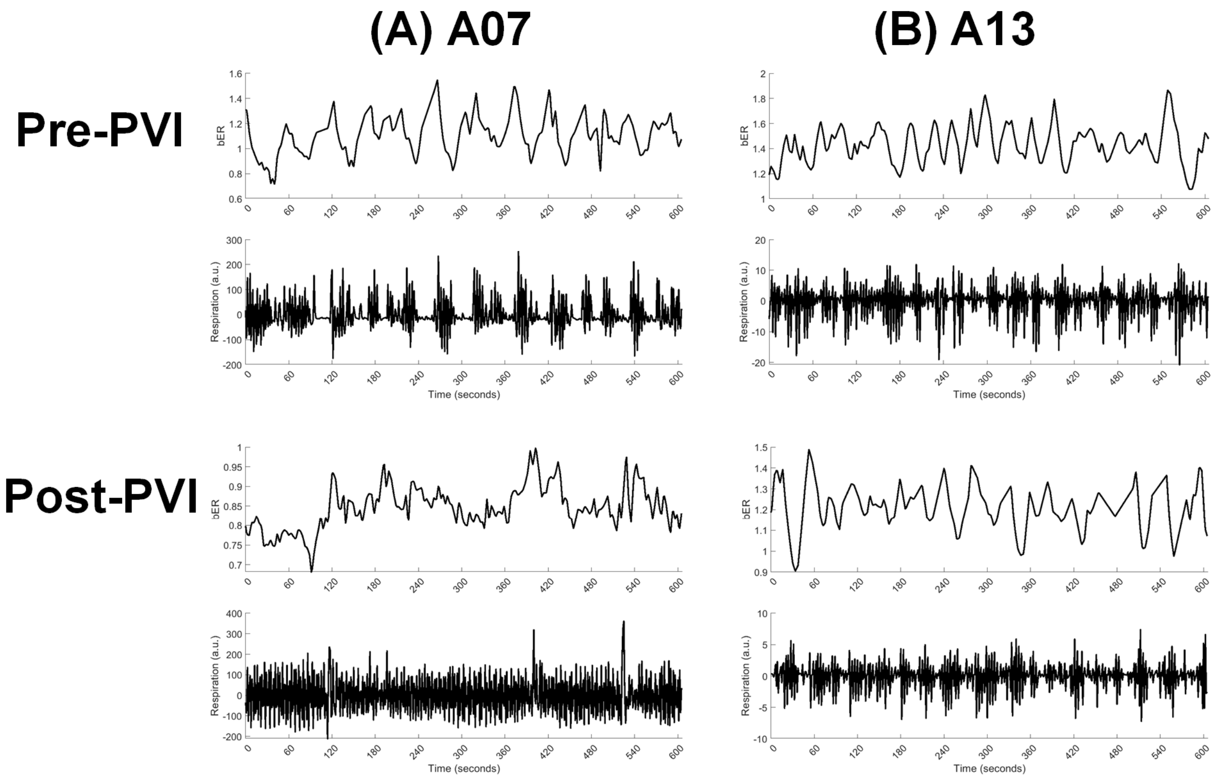


**Figure S2**. Time series of bER and respiration in two representative AF patients during spontaneous breathing at rest, before and after PVI. In the respiration time series, positive deflections above zero indicate inspiration, whereas negative deflections below zero indicate expiration. In patient A07, periodic breathing was evident in both the bER and respiration time series before PVI but was no longer observed after PVI. In contrast, patient A13 exhibited persistent periodic breathing both before and after PVI.

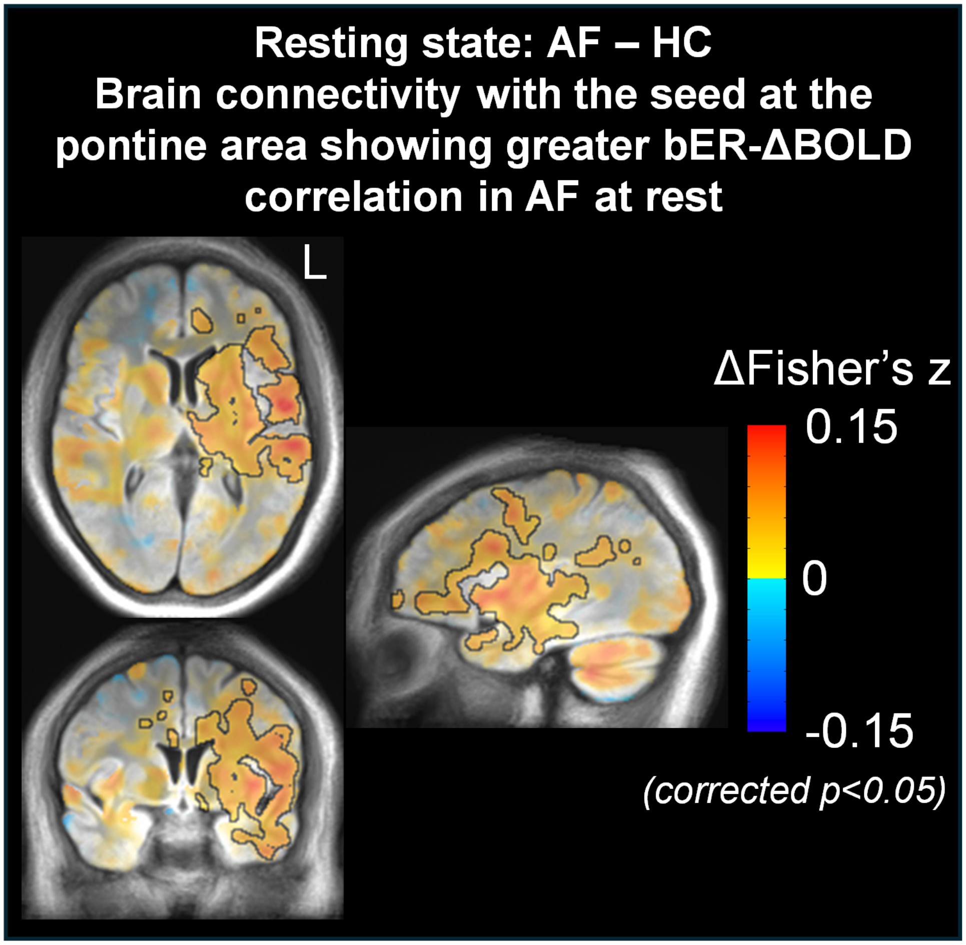


**Figure S3**. Comparison of resting-state functional connectivity between AF patients and HC, using a seed located in the pontine brainstem region that exhibited greater bER-ΔBOLD correlation in AF patients compared with HC. Connectivity between the pontine brainstem region and the left insula was significantly greater in AF patients than in HC. Warm colors indicate greater connectivity strength with the pons in AF patients relative to HC. Regions outlined in black achieved clustered-corrected $p<0.05$.

**Table S1**. Physiological metrics measured in AF patients and HC at rest

| Subject | HR (beats per minute) | | ToB (seconds) | | bER | | Periodic breathing |
|---|---|---|---|---|---|---|---|
| | Mean (SD) | CoV | Mean (SD) | CoV | Mean (SD) | CoV | |
| ***Patients with Atrial Fibrillation Before Pulmonary-Vein Isolation*** | | | | | | | |
| A01 | 65.77 (13.38) | 0.20 | 3.65 (0.59) | 0.16 | 1.00 (0.15) | 0.15 | 1 |
| A02 | 62.87 (8.42) | 0.13 | 4.70 (1.37) | 0.29 | 1.18 (0.07) | 0.06 | 0 |
| A03 | 78.86 (18.09) | 0.23 | 5.14 (2.42) | 0.47 | 1.00 (0.10) | 0.10 | 1 |
| A04 | 66.81 (13.19) | 0.20 | 7.01 (2.41) | 0.34 | 1.13 (0.10) | 0.08 | 0 |
| A05 | 88.33 (15.32) | 0.17 | 5.47 (1.88) | 0.34 | 1.30 (0.09) | 0.07 | 0 |
| A06 | 78.44 (14.89) | 0.19 | 7.20 (3.35) | 0.47 | 1.05 (0.10) | 0.09 | 0 |
| A07 | 43.72 (1.68) | 0.04 | 4.45 (2.23) | 0.50 | 1.09 (0.16) | 0.15 | 1 |
| A08 | 87.96 (15.23) | 0.17 | 6.92 (3.42) | 0.49 | 1.16 (0.12) | 0.10 | 0 |
| A09 | 68.41 (4.14) | 0.06 | 5.83 (4.09) | 0.70 | 1.19 (0.16) | 0.14 | 1 |
| A10 | 76.22 (11.66) | 0.15 | 9.05 (2.26) | 0.25 | 1.09 (0.11) | 0.10 | 0 |
| A11 | 98.84 (33.10) | 0.33 | 6.12 (1.86) | 0.30 | 1.28 (0.17) | 0.13 | 1 |
| A12 | 82.55 (17.40) | 0.21 | 2.85 (0.53) | 0.19 | 1.29 (0.09) | 0.07 | 1 |
| A13 | 97.08 (16.38) | 0.17 | 4.13 (1.16) | 0.28 | 1.43 (0.15) | 0.11 | 1 |
| A14 | 89.61 (17.29) | 0.19 | 4.78 (1.94) | 0.41 | 1.20 (0.11) | 0.09 | 1 |
| **Median [IQR]** | **77.53 [67.21-88.24]** | **0.18 [0.16-0.20]** | **5.52 [4.51-6.72]** | **0.37 [0.28-0.47]** | **1.17 [1.09-1.26]** | **0.10 [0.09-0.12]** | |
| ***Healthy Controls*** | | | | | | | |
| C01 | 55.13 (1.58) | 0.03 | 4.35 (1.82) | 0.42 | 1.18 (0.08) | 0.07 | 0 |
| C02 | 64.65 (3.86) | 0.06 | 5.11 (3.14) | 0.61 | 1.14 (0.08) | 0.07 | 1 |
| C03 | 46.51 (2.18) | 0.05 | 4.97 (0.68) | 0.14 | 1.08 (0.04) | 0.03 | 0 |
| C04 | 73.79 (2.76) | 0.04 | 6.66 (2.89) | 0.43 | 1.24 (0.09) | 0.07 | 0 |
| C05 | 69.90 (5.96) | 0.09 | 11.09 (3.94) | 0.36 | 1.25 (0.15) | 0.12 | 0 |
| C06 | 72.07 (6.28) | 0.09 | 4.31 (1.19) | 0.28 | 1.44 (0.12) | 0.08 | 0 |
| C07 | 72.27 (3.10) | 0.04 | 3.17 (0.33) | 0.10 | 0.96 (0.08) | 0.08 | 1 |
| C08 | 75.33 (1.72) | 0.02 | 5.05 (1.39) | 0.28 | 1.06 (0.20) | 0.19 | 0 |
| C09 | 85.11 (2.11) | 0.02 | 3.57 (0.27) | 0.08 | 0.78 (0.04) | 0.05 | 0 |
| C10 | 64.30 (2.63) | 0.04 | 3.52 (0.37) | 0.11 | 0.85 (0.05) | 0.06 | 0 |
| C11 | 74.28 (1.97) | 0.03 | 4.94 (1.25) | 0.25 | 1.30 (0.04) | 0.03 | 0 |
| C12 | 64.21 (2.18) | 0.03 | 4.09 (0.50) | 0.12 | 1.28 (0.07) | 0.06 | 0 |
| C13 | 59.17 (4.23) | 0.07 | 5.34 (2.28) | 0.43 | 1.08 (0.06) | 0.06 | 0 |
| C14 | 65.23 (2.48) | 0.04 | 5.53 (1.39) | 0.25 | 1.08 (0.04) | 0.04 | 1 |

| | | | | | | |
|---|---|---|---|---|---|---|
| **Median [IQR]** | **67.28 [64.23-73.41]** | **0.05 [0.03-0.06]** | **5.12 [4.15-5.28]** | **0.27 [0.13-0.40]** | **1.12 [1.06-1.25]** | **0.07 [0.05-0.08]** |
| ***Patients with Atrial Fibrillation Without Pulmonary-Vein Isolation vs. Healthy Controls*** | | | | | | |
| *p* | *0.027** | *<0.001** | *0.27* | *0.081* | *0.491* | *0.004** |

**p<0.05*

PVI, Pulmonary vein isolation; ToB, Time of breath; 0 represents absence of periodic breathing; 1 represents presence of periodic breathing.

**Table S2**. Physiological metrics measured at rest in AF patients before and after PVI.

| Subject | HR (beats per minute) | | ToB (seconds) | | bER | | Periodic breathing |
|---|---|---|---|---|---|---|---|
| | Mean (SD) | CoV | Mean (SD) | CoV | Mean (SD) | CoV | |
| ***Patients with Atrial Fibrillation Before Pulmonary-Vein Isolation*** | | | | | | | |
| A06 | 78.44 (14.89) | 0.19 | 7.20 (3.35) | 0.47 | 1.05 (0.10) | 0.09 | 0 |
| A07 | 43.72 (1.68) | 0.04 | 4.45 (2.23) | 0.50 | 1.09 (0.16) | 0.15 | 1 |
| A08 | 87.96 (15.23) | 0.17 | 6.92 (3.42) | 0.49 | 1.16 (0.12) | 0.10 | 0 |
| A11 | 98.84 (33.10) | 0.33 | 6.12 (1.86) | 0.30 | 1.28 (0.17) | 0.13 | 1 |
| A12 | 82.55 (17.40) | 0.21 | 2.85 (0.53) | 0.19 | 1.29 (0.09) | 0.07 | 1 |
| A13 | 97.08 (16.38) | 0.17 | 4.13 (1.16) | 0.28 | 1.43 (0.15) | 0.11 | 1 |
| A14 | 89.61 (17.29) | 0.19 | 4.78 (1.94) | 0.41 | 1.20 (0.11) | 0.09 | 1 |
| **Median [IQR]** | **82.60 [80.50-93.34]** | **0.19 [0.17-0.20]** | **5.21 [4.29-6.52]** | **0.38 [0.29-0.48]** | **1.21 [1.12-1.28]** | **0.11 [0.09-0.12]** | |
| ***Patients with Atrial Fibrillation After Pulmonary-Vein Isolation*** | | | | | | | |
| A06 | 52.88 (1.66) | 0.03 | 7.91 (5.15) | 0.65 | 1.05 (0.09) | 0.09 | 1 |
| A07 | 54.13 (1.34) | 0.02 | 3.28 (0.42) | 0.13 | 0.84 (0.06) | 0.07 | 0 |
| A08 | 52.38 (1.64) | 0.03 | 7.74 (6.75) | 0.87 | 1.06 (0.18) | 0.17 | 0 |
| A11 | 87.01 (1.20) | 0.01 | 6.09 (2.33) | 0.38 | 1.11 (0.05) | 0.05 | 0 |
| A12 | 62.68 (1.39) | 0.02 | 4.35 (1.08) | 0.25 | 1.18 (0.06) | 0.06 | 0 |
| A13 | 58.37 (3.02) | 0.05 | 4.99 (3.64) | 0.73 | 1.20 (0.11) | 0.09 | 1 |
| A14 | 64.37 (3.16) | 0.05 | 7.35 (2.75) | 0.37 | 1.15 (0.11) | 0.10 | 1 |
| **Median [IQR]** | **61.69 [53.50-63.53]** | **0.03 [0.02-0.04]** | **5.96 [4.67-7.54]** | **0.48 [0.31-0.69]** | **1.08 [1.05-1.16]** | **0.09 [0.06-0.10]** | |
| ***Patients with Atrial Fibrillation (After vs. Before Pulmonary-Vein Isolation)*** | | | | | | | |
| *p* | *0.031** | *0.016** | *0.219* | *0.219* | *0.016** | *0.219* | |

*$p<0.05$

PVI, Pulmonary vein isolation; ToB, Time of breath; 0 represents absence of periodic breathing; 1 represents presence of periodic breathing.
